# Improved resolution of influenza vaccination responses with high-throughput live virus microneutralisation

**DOI:** 10.1101/2024.09.13.24313549

**Authors:** Lorin Adams, Phoebe Stevenson-Leggett, Jia Le Lee, James Bazire, Giulia Dowgier, Agnieszka Hobbs, Chloë Roustan, Annabel Borg, Christine Carr, Silvia Innocentin, Louise MC Webb, Callie Smith, Philip Bawumia, Nicola Lewis, Nicola O’Reilly, Svend Kjaer, Michelle A Linterman, Ruth Harvey, Mary Y Wu, Edward J Carr

## Abstract

Influenza remains a significant threat to human and animal health. Assessing serological protection against influenza has relied upon haemagglutinin inhibition assays, which are used to gauge existing immune landscapes, seasonal vaccine decisions and in systems vaccinology studies. Here, we adapt our high-throughput live virus microneutralisation assay for SARS-CoV-2, benchmark against haemagglutinin inhibition assays, and report serological vaccine responsiveness in a cohort of older (>65yo) community dwelling adults (n=73), after the adjuvanted 2021-22 Northern Hemisphere quadrivalent vaccine. We performed both assays against all four viruses represented in the vaccine (A/Cambodia/H3N2/2020, A/H1pdm/Victoria/2570/2019, B/Yamagata/Phuket/2013, BVIC/Washington/02/201), using sera drawn on days 0 [range: d-28 to d0], 7 [d6-10] and 182 [d161-196] with respect to vaccination. We found population-level concordance between the two assays (Spearman’s correlation coefficient range 0.48-0.88; all P≤1.4 × 10^−5^). The improved granularity of microneutralisation was better able to estimate fold-changes of responses, and quantify the inhibitory effect of pre-existing antibody. Our high-throughput method offers an alternative approach to assess influenza-specific serological responses with improved resolution.

## Introduction

Influenza causes an estimated 389,000 deaths a year (1). Whilst annually updated multi-valent vaccines reduce this burden, better laboratory methods offering rapid, granular evaluation of the neutralising antibody response to these vaccines could further minimise this. Firstly, by facilitating refined yearly strain selection through high resolution quantification of the pre-existing antibody landscape. Secondly, by rapid, robust comparison between vaccine platforms, both existing (protein, adjuvanted protein, live-attenuated virus, or split virion), and upcoming (mRNA, and others). Thirdly, by providing a continuous response variable for systems vaccinology studies, to better leverage multi-modal high-dimensional and longitudinal datasets, themselves generated at significant cost and resource. Finally, by allowing prompt re-evaluation of human population immunity when an antigenically shifted influenza virus emerges, such as H5N1.

Currently, the haemagglutinin inhibition (HAI) assay is used to evaluate existing population-level immunity to guide annual vaccine strain selection, to confirm immunogenicity of these updated preparations, and as a correlate of protection. HAI assays are largely unchanged since first described in the 1940s (2,3): they are scalable, require minimal specialist laboratory equipment, but do not directly measure neutralising antibody, a critical effector arm of immunity. Virus will agglutinate red blood cells, naturally decorated with sialic acid, the ligand for influenza’s HA. Agglutination is inhibited by sera containing HA-specific antibodies. Live-virus neutralisation assays for influenza have existed for decades (4), but have to-date not been capable of displacing the HAI assay, largely due to limitations in the number of sera processed and their relative complexity.

## Results and Discussion

Here, we perform a head-to-head comparison of paired HAI, and a newly developed high-throughput live-virus microneutralisation (LV-N) assay. We demonstrate that large scale microneutralisation assays (see Materials and Methods) of 4 influenza strains are both feasible, and offer enhanced resolution. Adapting our SARS-CoV-2 approach, retains a similar throughput of ∼1,000-1,500 sera for 3 strains per week. We report sera from the AgeVAX study, an observational study of influenza vaccine responses in 73 community dwelling older adults (>65yo), with venepuncture on days 0 [range: d-28 to d0], 7 [d6-10] and 182 [d161-196] post-vaccination (**Table 1, Fig 1A**).

**Table 1.**
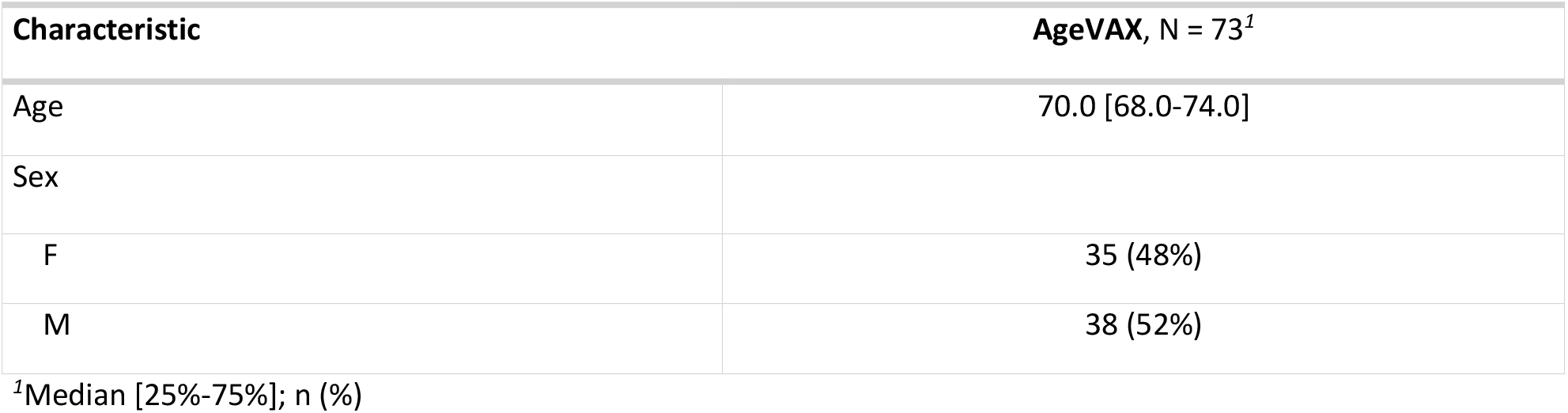

| Characteristic |  | AgeVAX, N = 73<sup>1</sup> |
| --- | --- | --- |
| Age |  | 70.0 [68.0-74.0] |
| Sex |  |  |
| F |  | 35 (48%) |
| M |  | 38 (52%) |

<sup>1</sup>Median [25%-75%]; n (%)

**Fig 1.**
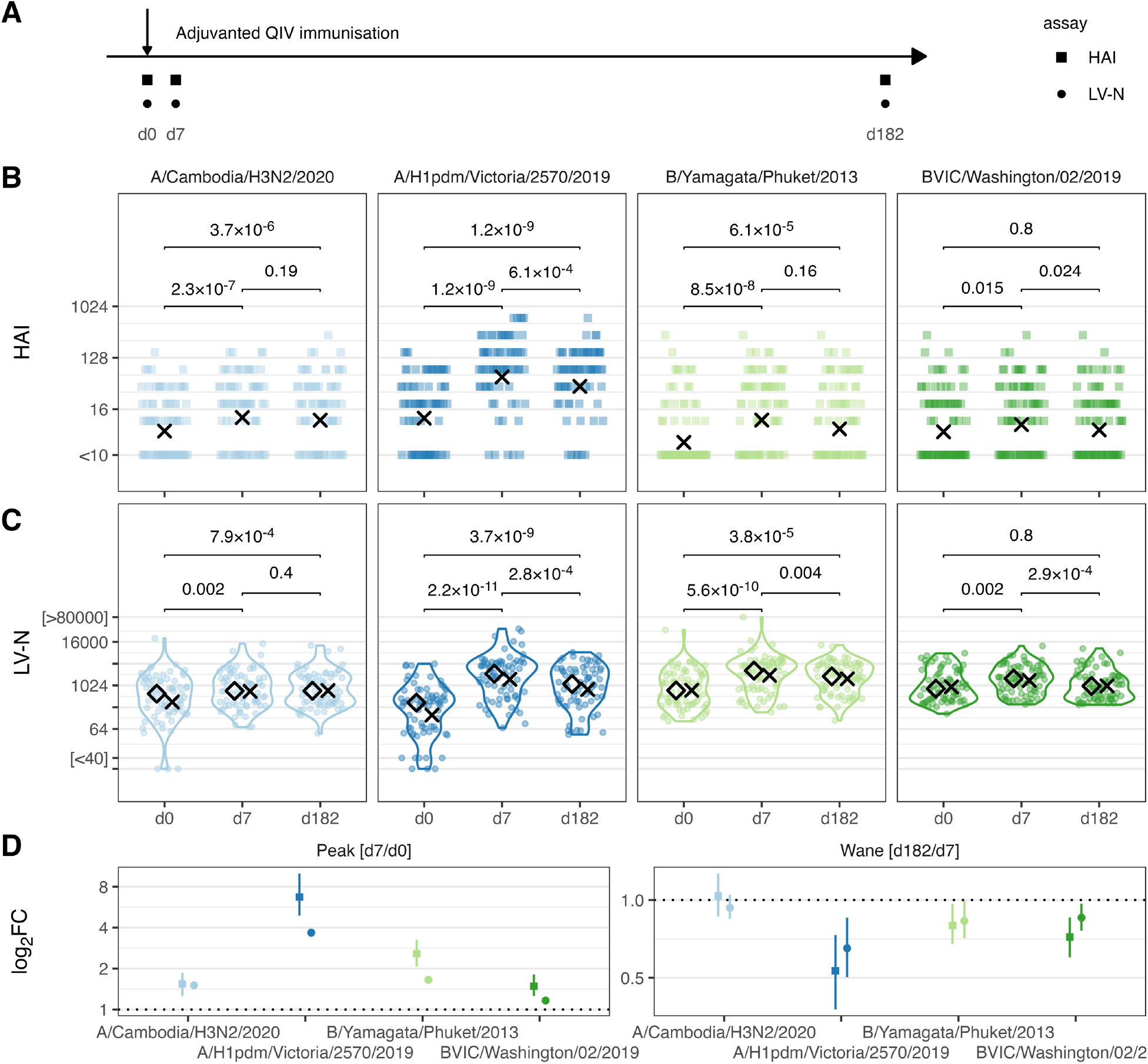
Serological vaccination responses quantified by haemagglutinin inhibition and high-throughput live influenza neutralisation assays in the AgeVax study. **(A)** AgeVax study design 73 individuals >65yo were vaccinated with adjuvanted quadrivalent influenza vaccine in the 2021-22 Northern Hemisphere season. **(B & C)** Haemagglutinin inhibition assay titres (HAI, **B**) or live-virus microneutralisation assay titres (LV-N, **C**), expressed as the reciprocal of the dilution at which 50% of virus infection is inhibited (IC_50_), at baseline (d0), and days 7 and 182 after vaccination (d7 & d182, respectively) for the flu viruses listed. **(D)** Log_2_ fold changes for peak (d7/d0) and wane (d182/d7) response as measured by HAI or LV-N (plotted as squares or circles respectively). Bootstrapped 95% confidence intervals are shown. In (B & C), crosses indicate geometric means, and *P* values from 2-tailed paired Wilcoxon signed rank tests are shown. In (C), diamonds indicate medians.

We first confirm increased antibody titres for all 4 influenza strains at d7 after administration of the 2021-22 vaccine using HAI (d7 vs d0 Wilcoxon *P*<0.015 for all strains, **Fig 1B**). Between paired sera, we find waning from d7 to d182 for two of the four immunised strains (A/Cambodia/H3N2/2020 *P*=0.19; A/H1pdm/Victoria/2570/2019 *P*=6.1×10^−4^; B/Yamagata/Phuket/2013 *P*=0.16; BVIC/Washington/02/2019 *P*=0.024, **Fig 1B**). We next repeated this immunogenicity assessment using our newly developed LV-N assay (**Fig 1C**). The boosting effect of vaccination is, once again, clear (d7 vs d0 *P*<0.002 for all strains, **Fig 1C**). With the LV-N assay, three of the four immunised strains showed significant waning between d7 and d182 A/Cambodia/H3N2/2020 *P*=0.4; A/H1pdm/Victoria/2570/2019 *P*=2.8×10^−4^; B/Yamagata/Phuket/2013 *P*=0.004; BVIC/Washington/02/2019 *P*=2.9×10^−4^. The quantitative nature of the LV-N assays offers improved resolution of the fold-change of the acute response (in the AgeVax study, the fold-change in titres d7/d0), with geometric mean fold-changes [95% confidence intervals] for LV-N estimating smaller reductions boosts with less uncertainty between these visits, when compared with HAI (**Fig 1D**). Furthermore, the waning (d182/d7) of B/Yamagata reached statistical significance only within the LV-N dataset (**Figs 1B & 1C**).

Thirdly, we plotted pairwise comparisons between HAI and LV-N for each strain at each of the three venepuncture visits (**Fig 2A**). Overall, the two assays showed a strong positive correlation according to their Spearman’s correlation coefficients. Considering differences between strains, B/Yamagata showed relatively weaker correlations than the other 3 strains. Next, considering differences over the 3 timepoints, the between-assay correlation for B/Victoria was weakest at d182, but no other strains showed marked differences over time.

**Fig 2.**
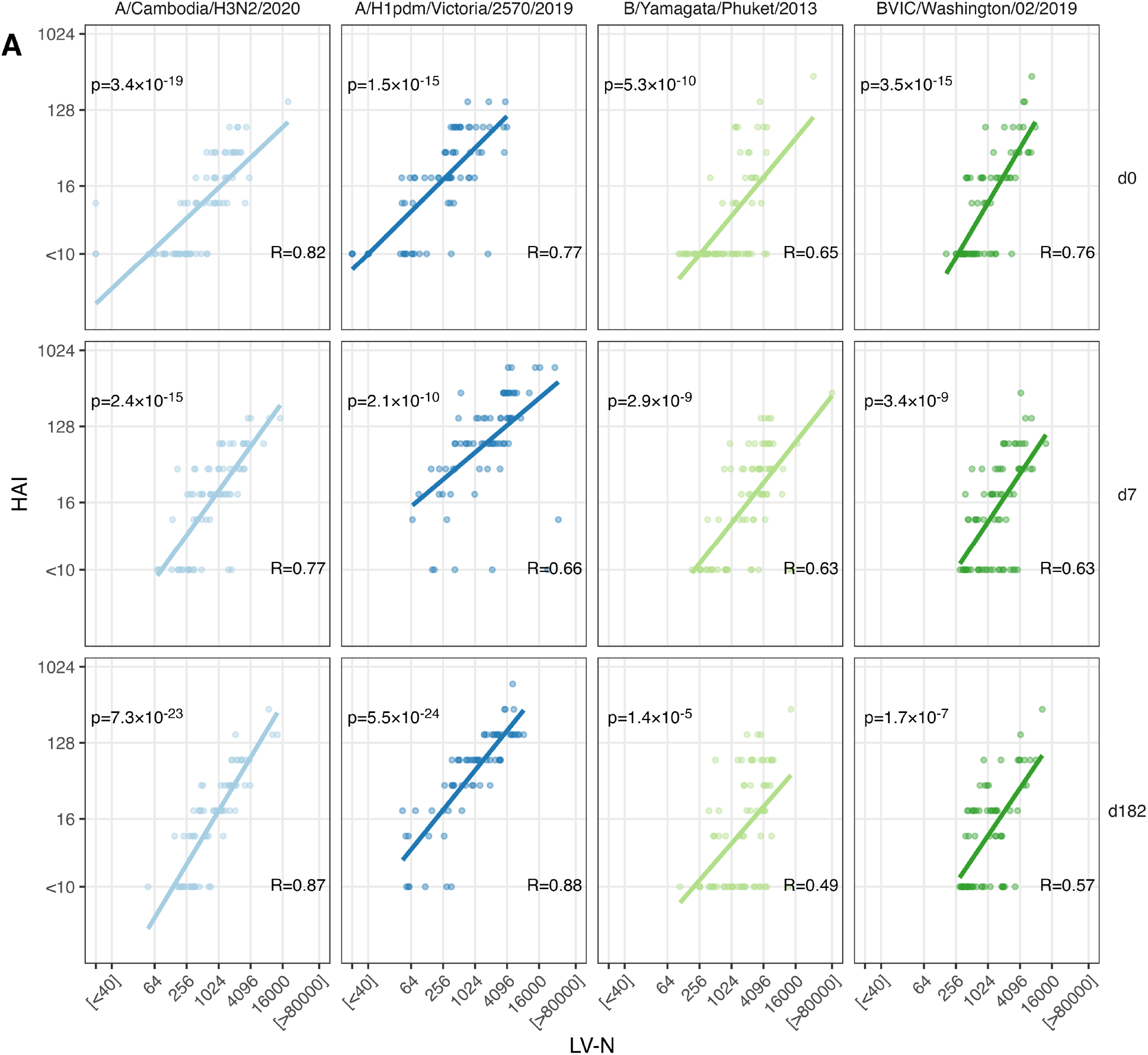
Comparison of haemagglutinin inhibition and high-throughput live influenza neutralisation assays. **(A)** Correlation between HAI and LV-N for each flu virus tested, at each venepuncture. In (A), Spearman correlation coefficients and *P* values are shown, and a fit line from linear regression plotted.

Despite the high concordance between assays at a population-level, we noted that for a given HAI titre, the range of corresponding LV-N results was large: for example, an HAI titre of 20 at baseline (d0) corresponded to an LV-N range of 43-1004 for influenza A/H1. This 23-fold difference suggests that immunologically important differences in vaccine responses are aggregrated into discrete HAI values. To address the question of whether LV-N offered improved sensitivity for biologically relevant heterogeneity over HAI, we returned to the observation that pre-existing antibody limits the fold-induction of antibody after vaccination (5).

Using HAI, we found that we could observe this negative relationship between higher baseline titres and lower fold-changes (d7/d0) for only FluA/H1pdm (P=0.029), with the other strains showing non-significant trends (**Fig 3A**). Repeating this comparison with LV-N, we found moderately strong negative correlations between pre-existing neutralising antibody and peak fold-change (d7/d0) for all variants (all P<0.009, **Fig 3B**). Coupling longitudinal LV-N-derived serological data with multi-modal datasets promises to unlock new potentials in systems vaccination, examining both peak and waned responses.

**Fig 3.**
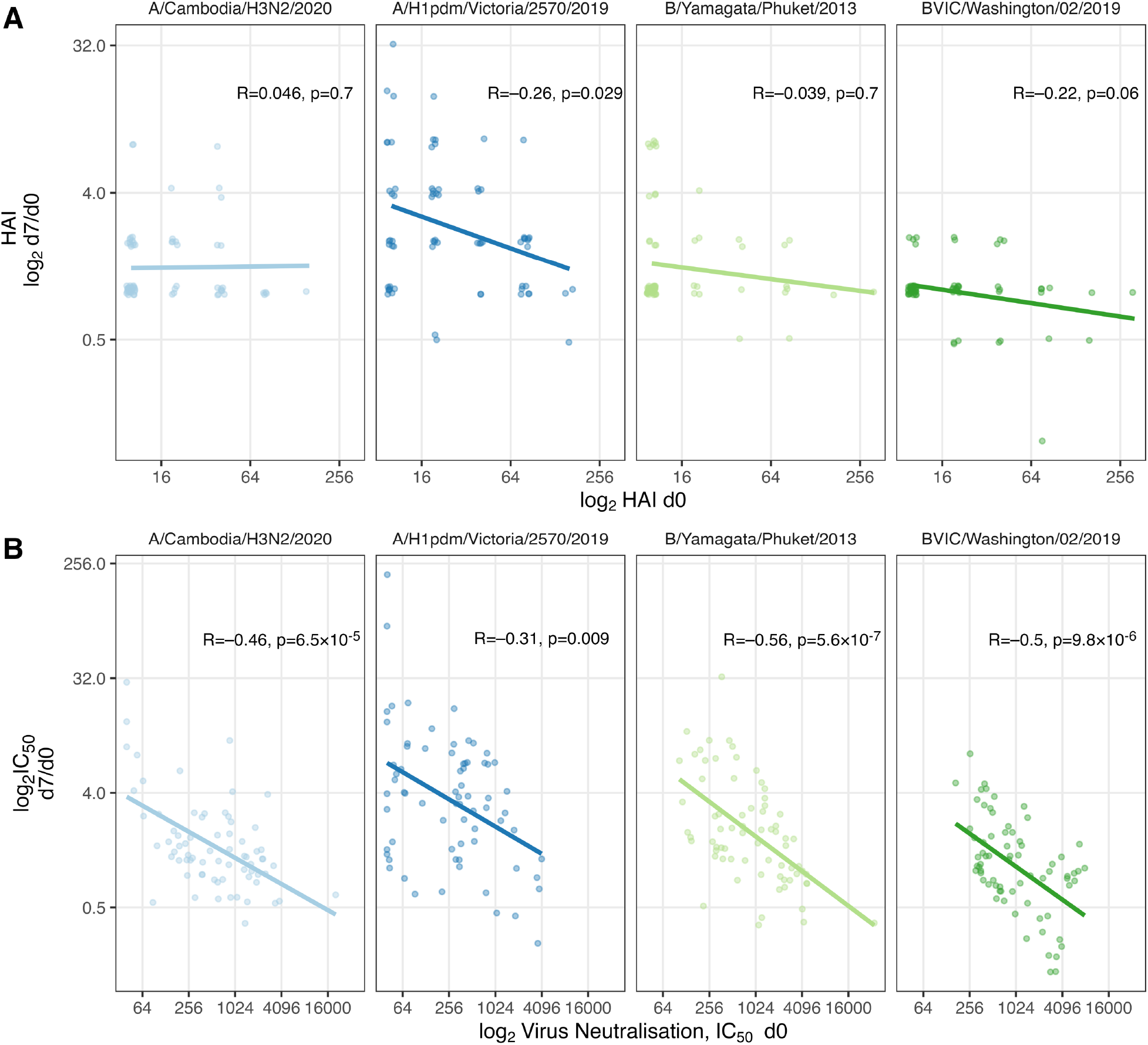
Pre-existing antibody inhibits subsequent vaccine responses. **(A & B)** Log_2_ fold changes of peak titres (d7/d0) plotted against d0 titres for HAI (**A**) or LV-N (**B**). In (A & B), Spearman correlation coefficients and *P* values are shown, and a fit line from linear regression plotted.

There are some limitations to our study. This single centre study enrolled older healthy adults, likely to mount a memory response, recalling both previously haemagglutinin-experienced B cells and recruiting naive B cells. It is plausible that a primary influenza vaccine response might be alternatively performant between these assays, particularly if significant IgM is produced, likely to perform well at HAI. Additionally, our live-virus microneutralisation approach reflects serological neutralisation of both HA and neuraminidase, whereas HAI does not assay NA-directed neutralisation. Other implementations of microneutralisation assays use alternative readouts for infection: for example, the WHO protocol uses an ELISA for nucleoprotein on fixed MDCK cells (reporting the lowest dilution at which the OD405 is below the midpoint of OD405 of virus-only and cell-only controls) (6), and a recently described next-generation sequencing assay testing infectivity/neutralisation of many DNA-barcoded viruses in parallel (7). Whilst, we have not directly compared with these methods, the DNA-barcoding approach can only find HA-related neutralisation, and the WHO neutralisation assay reports discrete rather than continuous data.

In summary, we have shown that high-throughput live-virus microneutralisation assays are non-inferior to HAI with strong correlations at population-levels. Further, we find that the continuous titre values returned by LV-N allow for finer dissection of vaccine responses, that are obfuscated by HAI. High-throughput influenza LV-N therefore has the potential to be a catalyst for rapid, robust assessment of existing antibody landscapes, new vaccine strain formulations, a step-change in systems vaccinology, and a facet of laboratory-based pandemic preparedness.

## Materials and Methods

### Study design, serum collection and ethics

The immune responses to vaccines in older persons study (AgeVax) enrolled community dwelling older adults already enrolled within the NIHR Cambridge BioResource to assess their response to seasonal influenza vaccine. AgeVax was approved by the Yorkshire & The Humber - South Yorkshire Research Ethics Committee (IRAS 277259, REC 20/YH/0101), and sponsored by the Babraham Institute, Cambridge. Healthy participants were enrolled if they were planning to receive the seasonal influenza vaccine as part of their routine care (in the UK, influenza vaccine is offered to all >65yo), were able to attend for consenting and venepuncture, and carried at least 1 allele of *HLADR*^*\**^*0701, HLADR*^*\**^*0401* or *HLADR*^*\**^*1101* (determined by single nucleotide polymorphism typing using the UK Biobank v2.1 Axiom array). Controlled comorbidities such as hypertension or hypercholesterolaemia were not exclusion criteria. Concomitant medications and comorbidities are not available for research purposes.

Venepuncture was performed up to 28 days pre-vaccination and on days 6-10, and 161-196 post vaccination. All participants received the adjuvanted quadrivalent influenza vaccine (Seqirus UK Ltd) via intramuscular injection. Blood was collected into silica-coated serum tubes and centrifuged at 2000rpm for 5 minutes and stored in aliquots at -80C. These sera were provided to the Francis Crick Institute for influenza assays described below.

### Cells and viruses

MDCK cells expressing the SIAT1 gene (MDCK-SIAT1) were maintained under selection in DMEM containing 1mg/ml Geneticin (G418) Sulphate (Stratech Scientific APE2513) and 1% Penicillin/Streptomycin (Sigma-Aldrich). Cells were seeded in DMEM containing 1% Penicillin/Streptomycin into 384-well cell culture microplates (Griener Bio-One Ltd) 18-20 hours prior to use in live-virus microneutralisation assays.

All influenza virus isolates used in this study were propagated in the allantoic cavity of 10-day-old embryonated hens’ eggs at 35°C for 48 hours.

### Hemagglutinin inhibition assays for influenza

Haemagglutination and haemagglutinin inhibition (HAI) assays were performed according to standard methods using suspensions of guinea pig RBCs (1.0% v/v) for A(H3N2) viruses and turkey RBCs (0.75% v/v) for A(H1N1)pdm09 and type B viruses with all serum samples pre-treated with receptor-destroying enzyme (RDE) from Vibrio cholera (8). Four haemagglutination units were used in all HAI assays. For A(H3N2) viruses, haemagglutination and HAI assays were conducted in the presence of 20 nM oseltamivir carboxylate (9).

### Live-virus microneutralisation assays for influenza

To perform high-throughput live virus microneutralisation assays for influenza we adapted our existing approach for SARS-CoV-2 neutralisation (10–12). MDCK-SIAT1 cells at 80% confluency were infected with selected influenza isolates in 384-well format, in the presence of 10-fold serial dilutions of participant serum samples, diluted in DMEM containing 1% Penicillin/Streptomycin. After 24 hours incubation at 37°C, cells were fixed using 4% formaldehyde (v/v), permeabilised with 0.2% TritonX-100 with 3% BSA in PBS (v/v), and stained for Influenza nucleoprotein (NP) using Biotin-labelled-clone-2-8C antibody produced in-house in conjunction with an Alexa488-Streptavidin (Invitrogen S32354) for influenza A isolates, or the mouse monoclonal B017 (Abcam, ab20711) in conjunction with an Alexa488 conjugated Goat anti-Mouse secondary (ThermoFisher, A21141) for influenza B isolates. Cellular DNA was stained using DAPI. Whole-well imaging at 5x magnification was carried out using an Opera Phenix (Perkin Elmer) and fluorescent areas calculated using the Phenix-associated software Harmony (Perkin Elmer). Virus inhibition by patient serum samples was estimated from the measured area of infected cells/total area occupied by all cells in each well and then expressed as percentage of maximal (virus only control wells). Infected cells were identified by presence of Influenza NP staining. The inhibitory profile of each serum sample was estimated by fitting a 4-parameter dose response curve executed in SciPy. Neutralising antibody titres are reported as the fold-dilution of serum required to inhibit 50% of viral replication (IC_50_), and are further annotated if they lie above the quantitative range (>40,000), below the quantitative range (<40) but still within the qualitative range (i.e. partial inhibition is observed but a dose-response curve cannot be fitted because it does not sufficiently span the IC_50_), or if they show no inhibition at all.

Viral isolates used in this study: A/Cambodia/e0826360/2020 H3N2, IVR-215 (A/Victoria/2570/2019) H1pdm, B/Phuket/3073/2013, B/Washington/02/2019.

### Data curation and analyses

HAI and live-virus microneutralisation titres were associated with anonymized metadata using R (v 4.2.2) and *tidyverse* (v 1.3.2) (13). Plots were generated using *ggplot2* (v 3.4.2), with *stat_summary()* to add geometric means or medians, as indicated in the figure legends. To plot HAI titres, sera with no inhibition were re-coded as 2.5. To plot LV-N titres, sera with either no inhibition of viral entry, or qualitative inhibition below the quantitative range (40-40,000), or inhibition greater than the quantitative range were re-coded as 5, 10 or 80,000 respectively. HAI and LV-N results were compared using two-tailed paired Wilcoxon tests, as implemented in the *rstatix* package, and resulting P values were plotted *ggpubr::geom_bracket()* and *ggtext::geom_richtext()*. Bias corrected 95% confidence intervals for fold-changes in HAI and LV-N were calculated from 1000 bootstraps generated with the *infer* package (14). For correlation between HAI and LV-N, Spearman’s correlation was used without censoring data above or below the quantitative ranges of HAI or LV-N. Anonymised data and R code are freely available via github:https://github.com/EdjCarr/AgeVax_HAI_LVN

## Data Availability

Anonymised data and R code are freely available via github.

https://github.com/EdjCarr/AgeVax_HAI_LVN

## Acknowledgements

The NIHR Cambridge Biomedical Research Center (BRC) is a partnership between Cambridge University Hospitals NHS Foundation Trust and the University of Cambridge, funded by the National Institute for Health Research (NIHR). We thank all NIHR Cambridge BRC volunteers for their participation and thank the NIHR Cambridge BRC staff for their contribution in coordinating the vaccinations and venepuncture. We would like to thank Marna Roos, Gita Mistry, Nicola Bex, Natasha Bowyer Irvine at the Francis Crick Institute and Heather Bath at the Babraham Institute. MAL is a Lister Institute Prize Fellow. EJC is supported by a Medical Research Council Clinician Scientist Fellowship (MR/X006751/1). This work was supported by the NIHR Cambridge BRC, the Dunhill Medical Trust (RPGF1910/223 to MAL), the Biotechnology and Biological Sciences Research Council (BBS/E/B/000C0427, BBS/E/B/000C0428, and the Campus Capability Core Grant to the Babraham Institute), the Medical Research Council (MR/X006751/1 to EJC), a UKRI Frontier award (EP/X022382/1 to MAL), in part funded by the NIH Centres of Excellence in Influenza Research and Response program Penn-CEIRR contract (75N93021C00015), and by the Francis Crick Institute which receives its core funding from Cancer Research UK (CC1114, CC2230, CC0102), the UK Medical Research Council (CC1114, CC2230, CC0102), and the Wellcome Trust (CC1114, CC2230, CC0102). The co-first and co-last authors contributed equally and are at liberty to list these authorships in any order on their curriculum vitae.

## Author contributions

LA: Conceptualization, Methodology, Investigation

PSL: Conceptualization, Methodology, Investigation

JLL: Project administration, Methodology, Investigation

JB: Methodology, Investigation

GD: Methodology, Investigation

AH: Methodology, Investigation

CR: Resources

AB: Resources

CC: Resources

SI: Methodology, Investigation

LMCW: Methodology, Investigation

CS: Resources

PB: Resources

NL: Resources, Supervision

NO’R: Resources, Supervision

SK: Resources, Supervision

MAL: Conceptualization, Funding acquisition, Supervision, Writing – review & editing

RH: Conceptualization, Funding acquisition, Investigation, Supervision, Writing – review & editing

MYW: Conceptualization, Funding acquisition, Investigation, Supervision, Writing – review & editing

EJC: Conceptualization, Data Curation, Formal Analysis, Visualisation, Writing – original draft, Writing – review & editing

